# The Functional Communication Checklist for People Living with Primary Progressive Aphasia: Study Protocol

**DOI:** 10.1101/2024.03.21.24304690

**Authors:** Jeanne Gallée, Jade Cartwright, Maya L Henry, Aimee Mooney, Brielle C Stark, Anna Volkmer, Connie Nakano, Rob J Fredericksen, Kimiko Domoto-Reilly, Paul K Crane

**Affiliations:** Department of Medicine, University of Washington, Seattle, WA, United States of America; School of Health Sciences, University of Tasmania, Launceston, Australia; Department of Speech, Language, and Hearing Sciences, University of Texas-Austin, Austin, TX, United States of America; Department of Neurology, Oregon Health & Science University, Portland, OR, United States of America; Department of Speech, Language, and Hearing Sciences, Indiana University, Bloomington, IN, United States of America; Division of Psychology and Language Sciences, University College London, London, United Kingdom; Department of Allergy and Infectious Diseases, University of Washington, Seattle, WA, United States of America; Department of Neurology, University of Washington, Seattle, WA, United States of America

**Keywords:** assessment, functional communication, primary progressive aphasia, person-centered

## Abstract

This study protocol describes the development of the first instrument of functional communication for people living with primary progressive aphasia (PPA), with future applications to other progressive conditions, with expert validation, item-level reliability analyses, and stakeholder input and outcomes. Progressive conditions like PPA require monitoring, and as such, re-assessment. Re-assessment poses the high risk of being burdensome, destructive, and of little use to the patient. As such, there is a significant need to establish a validated and reliable measure that (1) poses minimal patient burden and (2) captures communication ability in a strengths-based manner that is representative of daily communication needs and challenges. A strengths-based approach to assessment is widely recognized as the optimal way to promote patient autonomy, minimize harm, and implement functional treatment protocols and strategies. To date, there are no strengths-based assessment tools that were developed for people living with PPA. This study protocol describes our work to address this gap in clinical practice and research.

## 1 Introduction

Fifty thousand Americans are currently estimated to be living with primary progressive aphasia (PPA) [1], a clinical syndrome that initially presents with focal language decline and is typically attributable to pathological findings consistent with frontotemporal degeneration or Alzheimer’s disease (AD) [2–5]. People living with PPA (PwPPA) experience progressive decline in focal aspects of speech, language, and communication in the mild to moderate stages [2–5]. To date, there are three PPA variants established in the literature: the nonfluent/agrammatic, semantic, and logopenic with differentiated syndromic characteristics (see Table 1) [2–5].

**Table 1.**
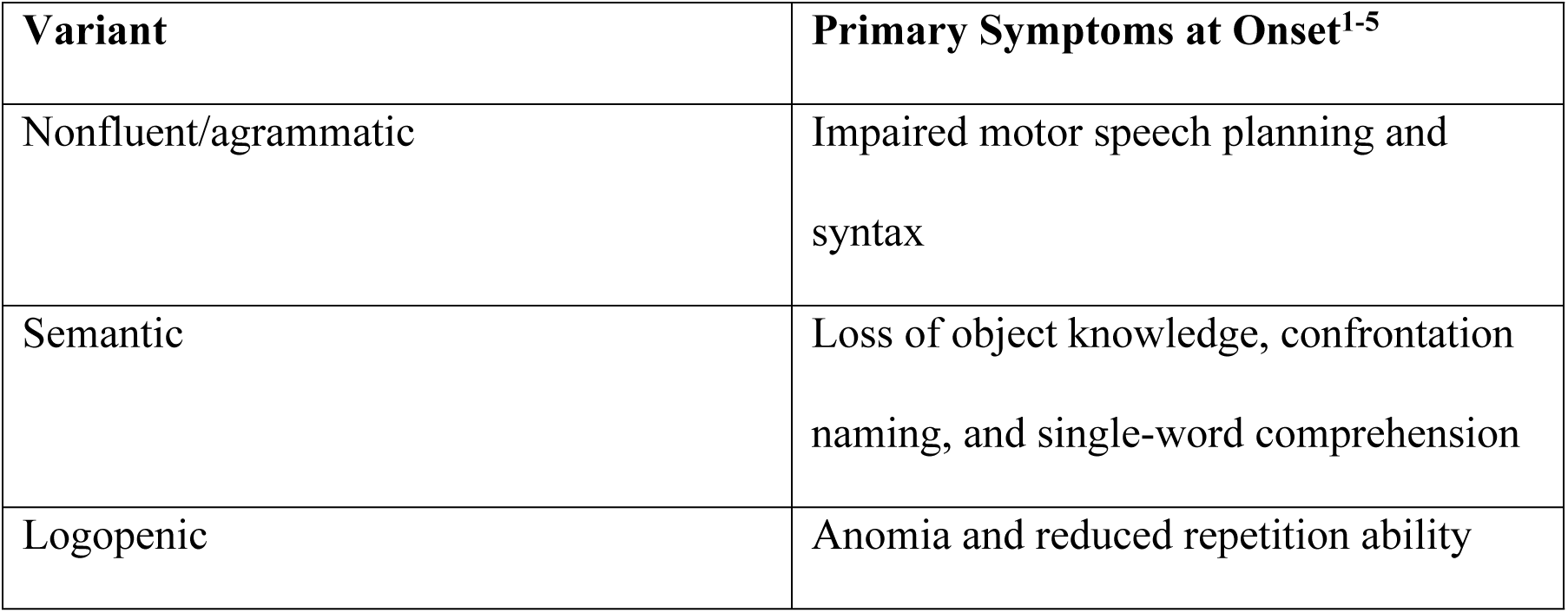
The Primary Symptoms of the Established PPA Variants.

To receive a PPA diagnosis, a person must experience prominent difficulty with speech and language at symptom onset that is the principal source of disrupted functioning and that is attributable to neurodegenerative disease [2–4]. PPA causes important changes in functional communication (FC), which is defined as the “transactional success” of expression [6], and which is a fundamental feature of human connection [7]. FC ability in PPA diverges by variant, individual differences, and time [5,8–10]. A person-centered approach—a holistic and humanistic method that promotes the autonomy and needs of the patient—is integral to establish how individual differences can impact a person’s success in FC [9,11] and to deliver individualized care [9,12].

PPA is a devastating, but relatively rare, condition [1]. As a result, a “gold standard” approach to care remains to be established [8]. Clinical assessment is essential to accurately identify and to formulate a specific diagnosis of PPA, and efficiently lead to treatment recommendations and post-diagnostic supports; however, to date, assessment procedures and documentation protocols are non-standardized or non-specific to this population [8,11]. These significant limitations have adverse effects on the accuracy and efficiency of diagnostic formulation and assessment outcomes, and result in irregularities in clinical evaluation protocols and cross-institutional characterization of research cohorts.

Particularly for rare neurodegenerative conditions, accurate and efficient assessment is critical to formulate a diagnosis, establish the effects of intervention, and monitor decline [8,11,13,14]. There is a need for direct assessment of functional communication (FC) in people with dementia [15] and PwPPA [16], particularly as this skill relates to early detection and designing optimal plans for intervention [13]. Early detection and treatment approaches also rely upon the identification of and distinction between the variant-specific impact on FC. Therefore, we have set out to develop a standardized checklist for FC, and in this paper present our protocol for this project. This study protocol centers on advancing assessment practices of FC for PPA, a critically important aspect of supporting activities of daily living, autonomy, and therapeutic intervention [7]. FC is interactive and contextual [7], yet traditional assessments of speech and language are not interactive and do not generalize to natural conversation [11,17]. As such, the current inventory of tools developed to capture and characterize FC decline in PPA is insufficient [8,16,17].

Therefore, the aim of this project is to create and validate an interactive tool to capture clinically relevant aspects of FC for people living any of the three variants of PPA (PwPPA). The primary outcome of this work will be the FC Checklist (FCC), a novel instrument to capture and track strengths-based change in FC ability. The FCC’s quantitative outcomes for speech, language, and communication performance create a common language that allows for cross-domain comparisons and consistency across evaluators and sites. The FCC will enable clinicians to quantify FC in a systematic and trackable manner and make appropriate and justified therapeutic recommendations [18]. The FCC will also serve as a research tool, providing more nuanced insight into the trajectory of a person’s cognitive-linguistic performance and impact on overall functioning, with the opportunity to provide participants with meaningful research outcomes. Moreover, clinicians and researchers will be able to use the FCC as a tool to provide valuable information to patients, care partners, and other providers to understand and actively engage with plans of care [14,19,20].

While some screening tools have been developed (e.g., the *Mini Linguistic State Examination*) [21], traditional aphasia instruments lack sensitivity to detect mild or early-stage PPA and fail to holistically evaluate FC ability [22]. Effective FC may be verbal, text-based, non-verbal, or multimodal [7]; however, few existing assessment tools examine non-verbal communication and instead focus on the other modalities in isolation. These tools thus fail to examine multimodal interaction—a crucial feature of day-to-day FC [22]. As such, there is a critical need for holistic, multimodal, and sensitive measures of FC ability, spanning clinical observation, quantification, and patient self-report [9,12].

We plan to develop a reliable tool that can be used over time and across institutions and providers. There is an important tool in this space, the Progressive Aphasia Severity Scale (PASS) [23]. The PASS was developed to capture decline across domains of speech, language, and communication in PPA. While the PASS provides a robust means of tracking change across an impressive range of linguistic domains, the PASS’s measurement of FC is restricted to a single item and is impairment-focused. Impairment-focused assessments are restrictive in that impairments are unreliable predictors of a person’s functional success [18,20,24]. In contrast, employing a strengths-based and person-centered approach reframes the patient as an active agent in their life [24] by capitalizing on their capabilities and their role in daily functioning [7,8,11,24,25]—a critical shift that is necessary to enhance the individualized and operationalized impact of clinical care. Finally, PwPPA report that most assessment protocols are time consuming and burdensome [11,14,25]. Minimizing assessment burden for people living with neurodegenerative conditions, such as PPA, is crucial to maintain trust and deliver respectful and patient-oriented care [11,14,25]. As such, there is a significant need for a person-centered, strengths-based, and minimally taxing measure of FC. We propose that the FCC will meet this need.

## 2 Materials and Methods

### 2.1 Study Objectives

1. <u>Tool Development</u>: curate an expert-validated clinical assessment tool of FC for PwPPA.

2. <u>Tool Implementation</u>: establish interrater reliability and validation of quantified scores of FC.

### 2.2 Ethics Approval

Approval for the study entitled “Assessment of Communicative Ability in Alzheimer’s Disease and Related Dementias” (STUDY00019344) was granted by the University of Washington’s Internal Review Board (IRB) on January 2^nd^, 2024.

### 2.3 Tool Development

#### 2.3.1 Participants

At least fifty speech-language pathologist (SLP) experts will be identified and recruited through the International SLT/P PPA Network (https://speechtherapyppa.builtbyknights.com/) as well as through a PubMed search for researchers with recent publications on FC in adults with neurodegenerative conditions.

#### 2.3.2 Experimental Approach

The purpose of the FCC is to broadly address whether specific features of speech, language, and nonverbal communication present as strengths or interferences in FC. To meet this purpose, item selection for the FCC will be guided by the methodological framework proposed by Kirshner & Guyatt (1985) [18]. As a conduit for appropriate clinical recommendations, which includes referral to speech and language services, the FCC must be an index that is (1) discriminatory [18], to distinguish people with and without FC challenges and (2) evaluative [18], to facilitate a level of sensitivity that captures change longitudinally in speech, language, and non-verbal communication. To ensure that the FCC meets the primary purpose in the context of this year-long award, we will use an electronic Delphi consensus process [26–28], consistent with the CREDES best practices [26] and technical recommendations [27,28] (see prototype generated in Phase One in Table 2 and Figure 1, Phases 3 and 4). The Delphi procedure is a structured technique to form consensus using collective intelligence [26–28]. To conform to standards, the procedure must be (1) anonymous, (2) able to actively engage a panel of experts, (3) iterative, and (4) able to provide feedback in the form of response summaries after each round [26–28]. The anonymity of web-enabled Delphi processes reduces pressure to conform to group opinion [26–28].

**Figure 1.**
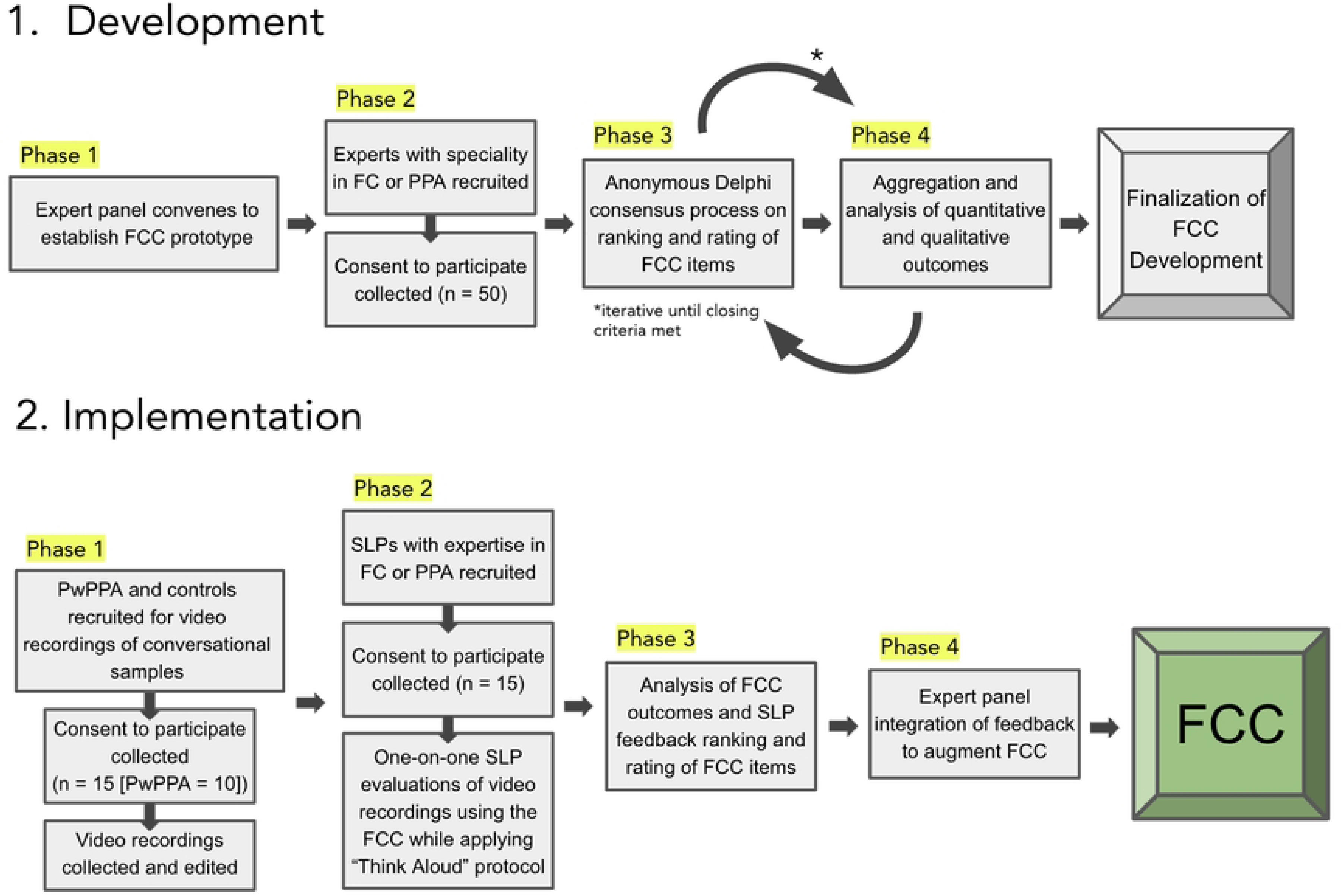
Study Flow.

**Table 2.**
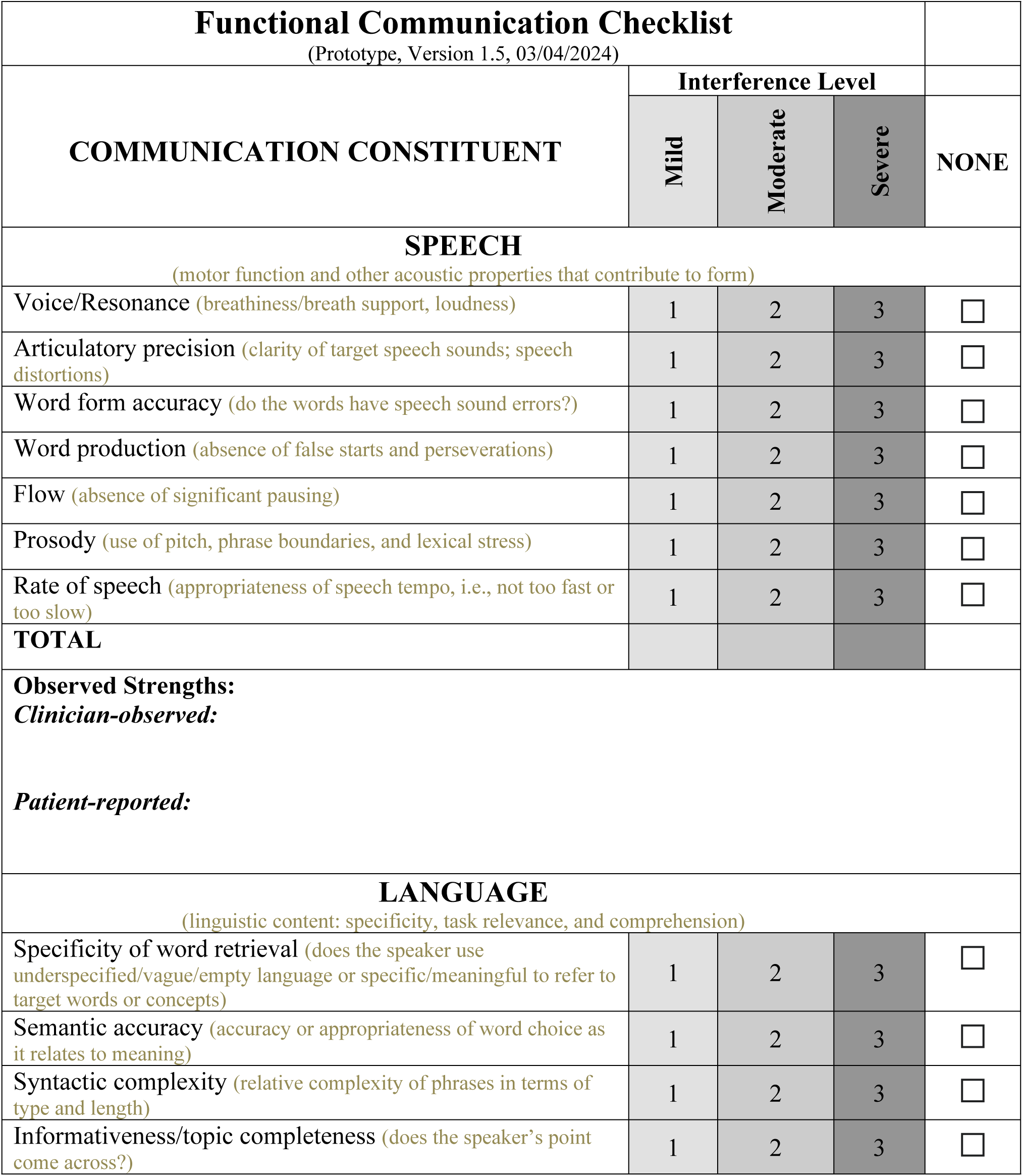
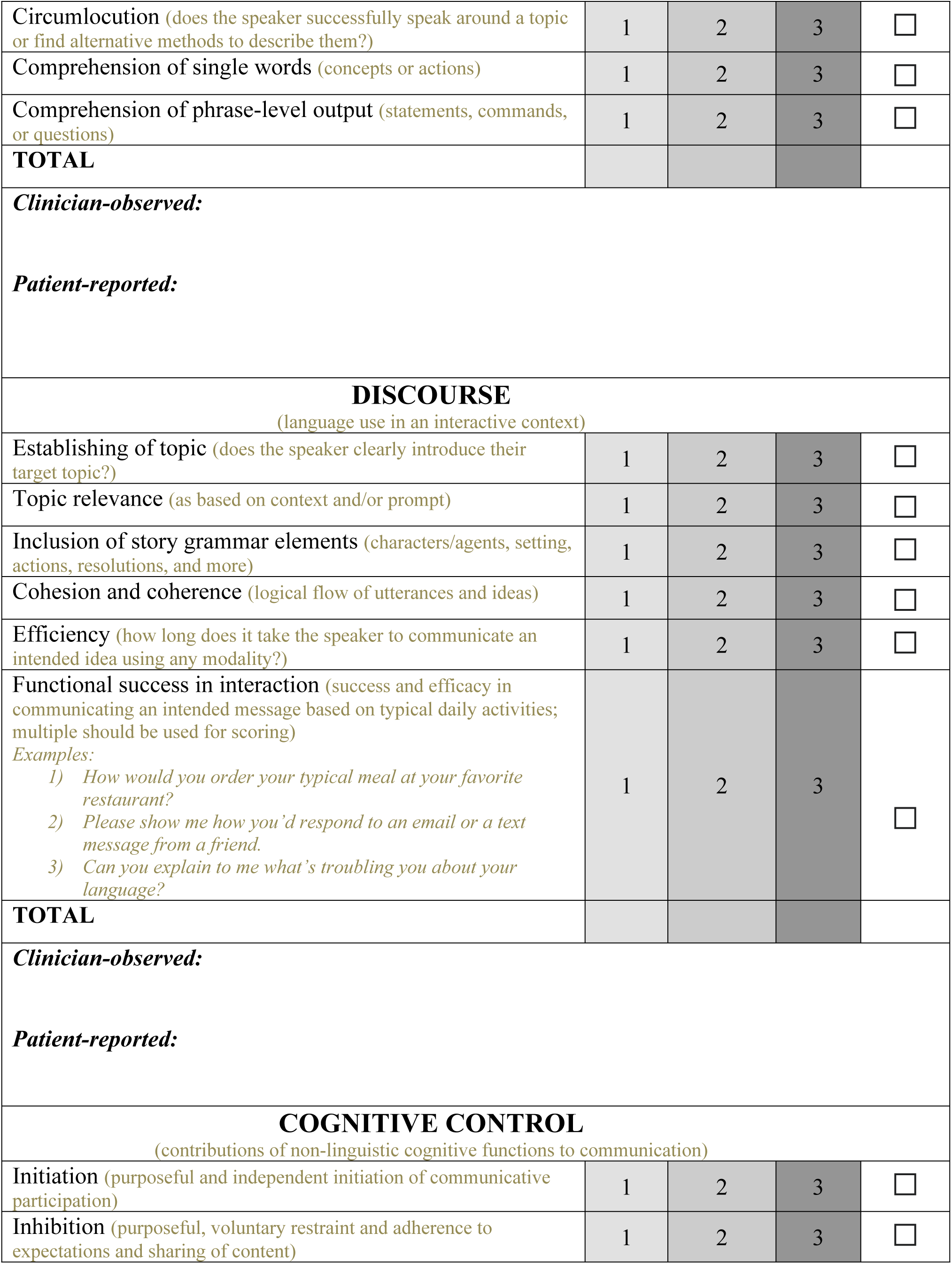
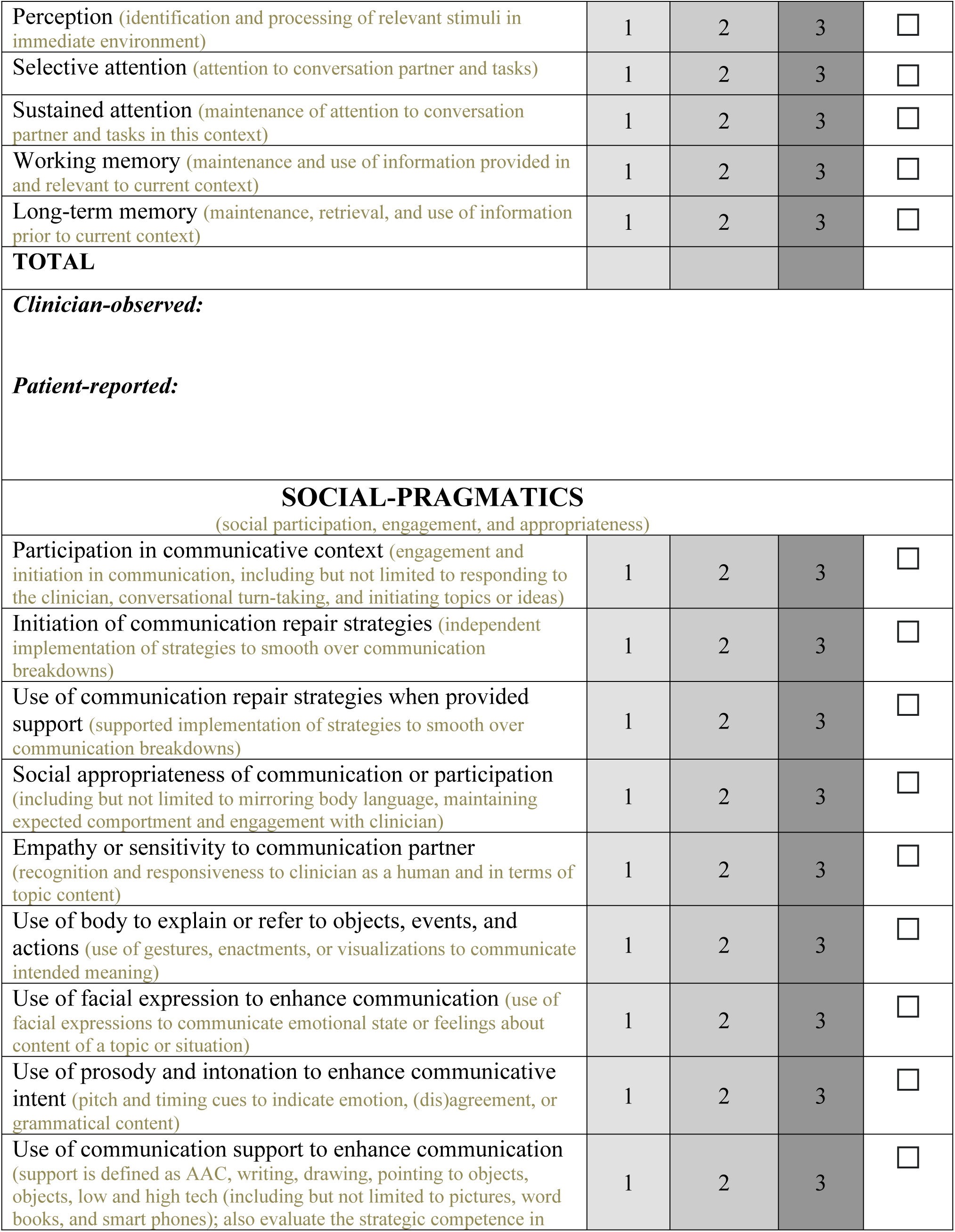
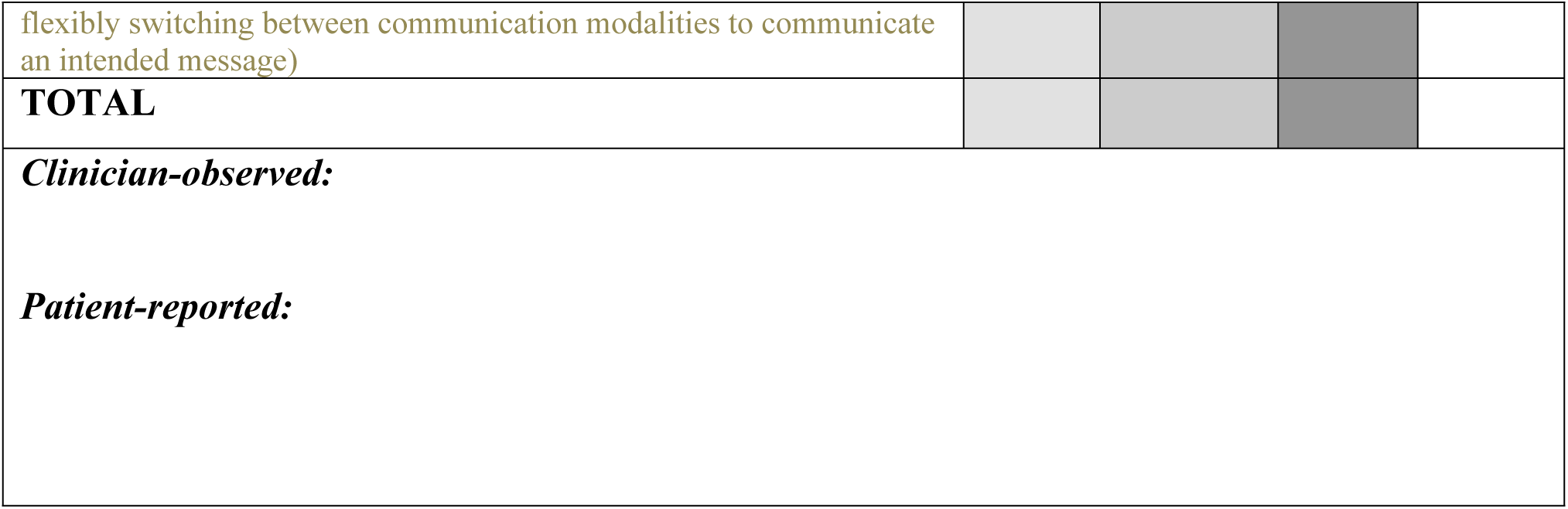
Prototype of the Functional Communication Checklist established in Phase 1 in preparation for the anonymous Delphi Consensus process to be conducted in Phases 2 through 4.

| Functional Communication Checklist |  |  |  |  |
| --- | --- | --- | --- | --- |
| (Prototype, Version 1.5, 03/04/2024) |  |  |  |  |
| COMMUNICATION CONSTITUENT | Interference Level |  |  | NONE |
|  | Mild | Moderate | Severe |  |
| **SPEECH**<br>(motor function and other acoustic properties that contribute to form) |  |  |  |  |
| Voice/Resonance (breathiness/breath support, loudness) | 1 | 2 | 3 | ☐ |
| Articulatory precision (clarity of target speech sounds; speech distortions) | 1 | 2 | 3 | ☐ |
| Word form accuracy (do the words have speech sound errors?) | 1 | 2 | 3 | ☐ |
| Word production (absence of false starts and perseverations) | 1 | 2 | 3 | ☐ |
| Flow (absence of significant pausing) | 1 | 2 | 3 | ☐ |
| Prosody (use of pitch, phrase boundaries, and lexical stress) | 1 | 2 | 3 | ☐ |
| Rate of speech (appropriateness of speech tempo, i.e., not too fast or too slow) | 1 | 2 | 3 | ☐ |
| **TOTAL** |  |  |  |  |
| **Observed Strengths:**<br>*Clinician-observed:*<br>*Patient-reported:* |  |  |  |  |
| **LANGUAGE**<br>(linguistic content: specificity, task relevance, and comprehension) |  |  |  |  |
| Specificity of word retrieval (does the speaker use underspecified/vague/empty language or specific/meaningful to refer to target words or concepts) | 1 | 2 | 3 | ☐ |
| Semantic accuracy (accuracy or appropriateness of word choice as it relates to meaning) | 1 | 2 | 3 | ☐ |
| Syntactic complexity (relative complexity of phrases in terms of type and length) | 1 | 2 | 3 | ☐ |
| Informativeness/topic completeness (does the speaker's point come across?) | 1 | 2 | 3 | ☐ |
| Circumlocution (does the speaker successfully speak around a topic or find alternative methods to describe them?) | 1 | 2 | 3 | ☐ |
| Comprehension of single words (concepts or actions) | 1 | 2 | 3 | ☐ |
| Comprehension of phrase-level output (statements, commands, or questions) | 1 | 2 | 3 | ☐ |
| **TOTAL** |  |  |  |  |
| **Clinician-observed:** |  |  |  |  |
| **Patient-reported:** |  |  |  |  |
| **DISCOURSE**<br>(language use in an interactive context) |  |  |  |  |
| Establishing of topic (does the speaker clearly introduce their target topic?) | 1 | 2 | 3 | ☐ |
| Topic relevance (as based on context and/or prompt) | 1 | 2 | 3 | ☐ |
| Inclusion of story grammar elements (characters/agents, setting, actions, resolutions, and more) | 1 | 2 | 3 | ☐ |
| Cohesion and coherence (logical flow of utterances and ideas) | 1 | 2 | 3 | ☐ |
| Efficiency (how long does it take the speaker to communicate an intended idea using any modality?) | 1 | 2 | 3 | ☐ |
| Functional success in interaction (success and efficacy in communicating an intended message based on typical daily activities; multiple should be used for scoring)<br>*Examples:*<br>1) How would you order your typical meal at your favorite restaurant?<br>2) Please show me how you'd respond to an email or a text message from a friend.<br>3) Can you explain to me what's troubling you about your language? | 1 | 2 | 3 | ☐ |
| **TOTAL** |  |  |  |  |
| **Clinician-observed:** |  |  |  |  |
| **Patient-reported:** |  |  |  |  |
| **COGNITIVE CONTROL**<br>(contributions of non-linguistic cognitive functions to communication) |  |  |  |  |
| Initiation (purposeful and independent initiation of communicative participation) | 1 | 2 | 3 | ☐ |
| Inhibition (purposeful, voluntary restraint and adherence to expectations and sharing of content) | 1 | 2 | 3 | ☐ |
| Perception (identification and processing of relevant stimuli in immediate environment) | 1 | 2 | 3 | ☐ |
| Selective attention (attention to conversation partner and tasks) | 1 | 2 | 3 | ☐ |
| Sustained attention (maintenance of attention to conversation partner and tasks in this context) | 1 | 2 | 3 | ☐ |
| Working memory (maintenance and use of information provided in and relevant to current context) | 1 | 2 | 3 | ☐ |
| Long-term memory (maintenance, retrieval, and use of information prior to current context) | 1 | 2 | 3 | ☐ |
| **TOTAL** |  |  |  |  |
| ***Clinician-observed:*** |  |  |  |  |
| ***Patient-reported:*** |  |  |  |  |
| **SOCIAL-PRAGMATICS**<br>(social participation, engagement, and appropriateness) |  |  |  |  |
| Participation in communicative context (engagement and initiation in communication, including but not limited to responding to the clinician, conversational turn-taking, and initiating topics or ideas) | 1 | 2 | 3 | ☐ |
| Initiation of communication repair strategies (independent implementation of strategies to smooth over communication breakdowns) | 1 | 2 | 3 | ☐ |
| Use of communication repair strategies when provided support (supported implementation of strategies to smooth over communication breakdowns) | 1 | 2 | 3 | ☐ |
| Social appropriateness of communication or participation (including but not limited to mirroring body language, maintaining expected comportment and engagement with clinician) | 1 | 2 | 3 | ☐ |
| Empathy or sensitivity to communication partner (recognition and responsiveness to clinician as a human and in terms of topic content) | 1 | 2 | 3 | ☐ |
| Use of body to explain or refer to objects, events, and actions (use of gestures, enactments, or visualizations to communicate intended meaning) | 1 | 2 | 3 | ☐ |
| Use of facial expression to enhance communication (use of facial expressions to communicate emotional state or feelings about content of a topic or situation) | 1 | 2 | 3 | ☐ |
| Use of prosody and intonation to enhance communicative intent (pitch and timing cues to indicate emotion, (dis)agreement, or grammatical content) | 1 | 2 | 3 | ☐ |
| Use of communication support to enhance communication (support is defined as AAC, writing, drawing, pointing to objects, objects, low and high tech (including but not limited to pictures, word books, and smart phones); also evaluate the strategic competence in | 1 | 2 | 3 | ☐ |
| flexibly switching between communication modalities to communicate an intended message) |  |  |  |  |
| **TOTAL** |  |  |  |  |
| ***Clinician-observed:*** |  |  |  |  |
| ***Patient-reported:*** |  |  |  |  |

Delphi processes have three distinct stages. The first is *conceptualization* [26], which entails defining the research goals, Delphi format, candidate FCC items, and additional questions for panelists. This stage (as described in Phase One, Tool Development in Figure 1) was conducted between February 21^st^ and March 10^th^, 2024 and carried out by the panel of PPA experts (JG, JC, MLH, AM, BCS, and AV). The second and third stages entail data collection and analysis (Phases 2 through 4, Tool Development in Figure 1). For this proposal, in Phase 2 of Tool Development, at least 50 PPA and/or FC experts from around the world will be invited to participate in iterative rounds of an online Qualtrics^XM^ [29] survey to establish rank order and rationales for the inclusion of each of the FCC items. Expert selection will be guided by the epistemological approach offered by Mauksch et al. (2020) [30], with a focus on panelist familiarity and expertise. In each round, participants will be asked to rank existing FCC items based on their clinical relevance and provide additional or alternative items to best evaluate FC in PPA. Participants will also be given the opportunity to explain their rationales and feedback for each FCC item. To minimize individual and collective bias, the survey introduction will draw explicit attention to possible biases [28]. Phases 3 and 4 of Tool Development will occur iteratively until closing criteria have been met (see below).

#### 2.3.3 Statistical Analysis

We will implement a mixed-methods approach of qualitative and quantitative analyses to evaluate panelist feedback. Panelists will receive aggregate descriptive outcomes of rank order and general inclusionary/exclusionary rationale after each round, as well as summaries of qualitative suggestions for additional or alternative items. This feedback will explain item ranking per FCC constituent and the resulting modification of the checklist items. Closing criteria will consist of a minimum of 80% consensus for inclusion of FCC items [26–28] with statistical stability (insignificant difference in item consensus) over the minimum of four rounds [26] of the survey. Following the final round of the Delphi procedure, panelists will receive the comprehensive statistical analyses and results. Investigator JG and Co-Investigators PKC and RJF will code the qualitative feedback by content type (e.g., rank order rationale, rationale for proposed alternative/revised item, etc.) using Dedoose qualitative coding software [31] with acceptable interrater reliability of 80% or higher [28].

We will then utilize a qualitative “memo-ing” process based on the Grounded Theory model of qualitative inquiry [32] to summarize themes within each content type. Investigator JG and Co-Investigators PKC and RJF will independently summarize themes and subsequently meet to discuss and reconcile differences in interpretation and achieve consensus. For descriptive statistics, arithmetic mean values and standard deviations will be calculated. Interquartile ranges will be used to assess consensus. Bipolarity analyses will be conducted to examine whether there are sub-group differences despite in-group consensus. Finally, outlier analyses will be conducted to detect whether there are differential interpretations of certain items due to statement comprehensibility or other reasons revealed in the qualitative feedback.

### 2.4 Tool Implementation

#### 2.4.1 Participants

##### 2.4.1.1 SLPs

After the FCC has been finalized by meeting closing criteria of the Delphi procedure, 15 SLPs who were not involved in developing the FCC and who have specialization in PPA will be recruited to pilot the checklist to rate FC based on the discourse samples from video recordings described below (see section 2.4.1.2 Video Curation). Expert SLPs will be recruited through the International SLT/P PPA Network [33], which has a reach of upwards of a hundred of relevant experts available to be recruited for this purpose. Additionally, non-expert SLPs who work with adult populations will be recruited through online forums and the University of Washington. Participants will receive a one-time payment of $100 for their participation in Phase 2 of Tool Implementation.

##### 2.4.1.2 PwPPA

We will recruit individuals that span the PPA spectrum, with three to four examples of each of the three established variants [2], including heterogeneous profiles [4,5], and five age-matched controls with typical cognition. The purpose of the control group is to anchor the typical range of variability of communication. Participants will be recruited from the University of Washington’s Alzheimer’s Disease Reach Center (UW ADRC) and affiliated Memory and Brain Wellness Center. Numbers of unimpaired participants are surpassed by the active cohorts of the UW ADRC’s Clinical Core and Registry. Eligible and interested participants will be consented to participate in an identifiable video recording. Participants will receive a one-time payment of $150 for their participation in the assessment described in Phase 1, Tool Implementation.

#### 2.4.2 Experimental Approach

##### 2.4.2.1 Video Curation

Video recordings will be collected of PwPPA and controls using a procedure consistent with the validation process described for the *Clinical Dementia Rating* (CDR®), a global staging scale of individual domains [34,35]. The assessments will consist of three naturalistic discourse samples, elicited by a conversational in-take to establish the PwPPA’s self-described strengths and needs, and one closed-ended and one open-ended discourse prompt prior to completing the *Quick Aphasia Battery* (QAB) [36] to establish performance across domains of articulation, auditory comprehension, lexical retrieval, motor function, reading, repetition, and semantic processing. Patient-reported outcome measures will be gathered by asking PwPPA to rate the relative difficulty of the discourse sample tasks using an aphasia-friendly, stakeholder-approved 5-point visual scale (0 = high burden, 4 = no burden) from the freely available, psychometrically evaluated *Aphasia Impact Questionnaire-concise* (AIQ-concise) [37]. The AIQ-concise scale offers a selection of gender and race visualizations to allow PwPPA to choose the pictorial representations that closest align with their visualizations of self. Patient ratings of perceived task-burden for the discourse tasks will be compared to those of the comprehensive QAB to establish whether discourse-based tasks are perceived as less burdensome.

##### 2.4.2.2 FCC Implementation

In separate 45-minute online Zoom calls, the 15 SLPs will watch the participant responses to the open and closed-ended discourse prompts. Following each case example, the SLPs will be asked to fill out the FCC via a Qualtrics^XM^ [29] poll. SLPs will be asked to simultaneously record their thought processes as they carefully consider the applicability of each FCC item, consistent with the “Think Aloud” protocol [32,38,39]. Application of the “Think Aloud” protocol will result in the collection of qualitative targeted thinking to further refine the FCC. Two additional questions will be asked: (1) How effective is this person’s communication (1 = very effective, 5 = acceptable, 10 = ineffective) and (2) Rate the level of impairment of FC (0 = typical, 0.5 = questionable, 1 = mild, 2 = moderate, 3 = severe). The latter question maps onto the PASS’s “Functional Communication” item [23], whereas the former allows for a “big picture” rating and how this relates to the survey’s other items. Following the completion of these Zoom calls, results will be analyzed for item-level inter-rater reliability. Agreement between the participant samples and “gold standard” ratings for each case example, generated by the study team and expert panelist consensus, will also be analyzed.

We will then gather targeted feedback for the people assessed, providing the person and people who want to communicate with them specific and evidence-based recommendations to address areas of concern and improve FC. To generate this targeted feedback [9,11,12,25] based on the FCC’s structure, the study team will develop informational guidance and clinical recommendations for each of the behaviors included. The purpose is multifold and intended for patients, care partners, researchers, and clinicians. In the context of a Zoom-based focus group, study team members will generate concrete descriptions and guidance to address the possible interference posed by each behavior described in the FCC (e.g., “word form” or “rate”, see Table 2) accompanied by publicly available and aphasia-friendly visuals [40]. Consistent with the best practice principles for PPA [8] and expert recommendations, the feedback template will explain the purpose of the discourse tasks collected in Phase 1, Tool Implementation, contextualize the communication behavior outcomes, and provide tailored recommendations to enhance FC. The feedback template will be made publicly available online through the UW ADRC website for anyone to input FCC outcomes and receive individualized guidance for strengths and relative interferences.

#### 2.4.3 Statistical Analysis

These analyses are exploratory, with the goals of (1) understanding the assessment items so that a future grant proposal can make any necessary improvements, and (2) generating hypotheses for that project. To assess inter-rater item reliability, we will use the version of the kappa statistic that allows for different sets of raters. Thresholds for acceptable kappa values vary, but we will consider a kappa of 0.60 to be usable [41]. We are also aware of the “kappa paradox” that can arrive if a rater has low sensitivity or specificity. Future work may involve forming summary measures for the subdomains once items have been finalized. Qualitative feedback from the “Think Aloud” procedure will be coded using Dedoose qualitative analytic software [31] by study team members trained in qualitative analysis. This round of coding will apply a fixed code for each FCC item discussed. Then, using the Grounded Theory-based “memo-ing” process [32], three analysts will each independently review the feedback associated with each FCC item, and draft a memo outlining issues/themes/concerns for each. Analysts will subsequently meet to share and reconcile differences in interpretation and achieve consensus regarding thematic content at the item level. The resulting final, integrated memo will inform finalization of the FCC. The FCC is a formative measure rather than assessing a latent trait, so a weighted score may or may not be advisable, depending on how well the items correlate with each other and with overall function; for certain individuals, strength in one item may be a prominent feature of FC, independent of the presence or absence of other strengths.

### 2.5 Protection of Human Participants

#### 2.5.1 Participants of Phase 1, Tool Implementation

Our goal for the proposed research is to recruit and enroll 9-10 individuals with a diagnosis of PPA. The majority of individuals evaluated annually as part of the Clinical Core program at the UW ADRC have also consented to being approached for additional research studies. Participants eligible for the study will be patients who have received a diagnosis of PPA by a neurologist (KDR), enrolled in the ADRC Clinical Core or UW Memory and Brain Wellness Registry, and are able to comply with the experimental protocol. Patients who cannot comply with the experimental protocol due to hearing, English proficiency, vision, or cognitive impairment will be excluded. Similarly, patients who do not consent to being video-recorded and having these recordings shared on UW Sharepoint for educational purposes, accessible through the UW ADRC website, will also be excluded.

Participants who respond positively to recruitment will be given a full explanation of the project by study staff. Per NIH policy, as a part of the informed consent process, we will also collect contact and demographic information from each of our PPA and control participants. Informed consent will be obtained from all people participating in this study, and all methods of recruitment and experimental protocols will be approved by the institutional review board of the University of Washington. Consent will be obtained from the participant or their legal representative, where the participant would at a minimum provide assent. A copy of the signed consent form will be provided to all enrollees.

#### 2.5.2 Participants of Phases 3 and 4, Tool Development and Phase 2, Tool Implementation

Our goal is to also recruit at least 65 SLPs with either expertise in PPA or general knowledge in the assessment and care of adult populations. These SLPs will be recruited through the University of Washington, national and international working groups, and through online forums geared towards this target population. Exclusionary criteria will include inability to commit to the time required for the experimental protocols for an online survey (Phases 3 and 4, Tool Development) as well as dissent to being recorded for those participating in Zoom videoconferencing calls (Phase 2, Tool Implementation). The participant will be recruited through advertisement materials posted online and through physical flyers. Each eligible participant will be provided with information describing the purpose of the project, the experimental procedures, potential risks, and benefits, and required time commitment. If the participant would like to participate, they will receive an email to with an attachment for the informed consent documentation. Each participant will receive a copy of the signed informed consent document, and the original will be retained by the PI and stored on a secured Drive in Co-Investigator KDR’s lab.

#### 2.5.3 Protection of Participant Data

Participant information as well as behavioral data will be collected according to the procedures described within **Materials and Methods**. The data will consist of clinical assessments as well as acoustic signals and digital video recorded during study visits. Participant information, such as participant histories and clinical assessments, will be recorded by entering the data directly into an electronic data capture system. The system used will be a secure, HIPAA compliant implementation of the REDCap Research electronic data capture software, hosted by the UW ADRC. This web-based data capture system is designed specifically for human participants research, and is used by over 1,000 institutions worldwide. It provides audit trails for tracking data manipulation and user activity. Access is controlled by a secure web authentication system and SSL encryption, and will be limited to the PI, Co-Investigators, and other IRB-approved study staff only. Behavioral language data will be stored here. The digital video recorded during the study will be stored on a secure, password-protected drive hosted by the UW Sharepoint. Access to study data on this drive will be limited to approved and verified individuals. Paper records will be accessible only to the Co-Investigator, PI and IRB-approved study staff. Participant-identifiable information such as the master list matching participant names to ID numbers will be stored for 5 years and then destroyed. To maintain confidentiality of the participants and their records, participants will be identified in all study records and computer files by a three-digit sequential numeric code.

The master list matching participant names to ID numbers will be stored in a password-protected and encrypted digital file that is only accessible to study staff. The privacy and confidentiality of participants participating in this study will be protected at all times. Study procedures, including the explanation of the study and informed consent process, will take place in a private office space. Participants will be referred to throughout study files only by an anonymized numerical code. All computer files containing participant-identifiable information will be kept in secure, access-restricted, encrypted digital storage; any physical files pertaining to study participants will be kept locked in a file cabinet in the Co-Investigator’s (KDR) lab. Only IRB-approved study staff will have access to review study records. No sensitive information will be collected during this study that would require reporting to state or local authorities.

#### 2.5.4 Potential Risks

The potential risks to participants from participating in this research are minimal. All participants participating in Phase 1 of Tool Implementation will complete behavioral assessments at a single timepoint. However, the extent of potential fatigue is not beyond what may be experienced in any other daily activity. There is a potential risk for discomfort due to the physical environment of being tested in a private room while being recorded. To preempt this, all participants will receive ample transition time to the space to help them get comfortable and to prepare them for the actual assessment.

#### 2.5.5 Protections Against Risks

All information about the participants will be kept confidential. To maintain confidentiality of the participants and their records, each participant will be assigned an identification number and referred to by this number. The master list matching participant names to ID numbers will be stored in a locked cabinet that is only accessible to the PI and Co-Sponsor. Only IRB-approved study staff will have access to the data. The records will be kept for approximately 5 years after completion of the study. This period will be needed to verify results prior to publication. The PI will be responsible for applying for and maintaining full IRB approval. In addition, all project personnel will be required to complete human participants training. The University of Washington’s Federal-Wide Assurance requires that all University of Washington research with human participants, regardless of funding source, abide by the Belmont principles of respect, beneficence, and justice and the federal regulations in 45 CFR 46. Further, it states that University of Washington will provide initial and continuing education to personnel conducting research with human participants to help ensure that these ethical standards are met. To assist in this process, University of Washington has subscribed to the Consortium for IRB Training Initiative in Human Subjects Protections (CITI).

To be certified for human participant research, key project personnel must complete the CITI tutorial every three years; this training must be supplemented annually through CITI refresher tutorials or through attendance at one or more educational sessions held by the IRB. Records verifying the completion of the above training for all individuals will be maintained by Co-Investigators KDR and PKC. Participants in Phase 1 of Tool Implementation will be closely monitored by study staff during the study for fatigue, discomfort, or any other adverse events. The PI will be responsible for the reporting of any adverse events that occur over the course of the study. Adverse event reporting will be done according to the guidelines of the University of Washington’s Human Subjects Division and our IRB. All major and minor adverse events will be reported to the IRB. Should a participant express or show signs of discomfort or fatigue that cannot be resolved with short periods of rest, the protocol will be terminated. No special precautions are required before, during or after the study by the participant.

#### 2.5.6 Potential Benefits of the Proposed Research to Human Participants and Others

For participants with PPA, we will provide information regarding the results of all testing at the individual’s request. This information may be useful for documenting symptom progression and further explanation of their impact on daily participation and communication.

## 3 Results

This study was approved for funding from the University of Washington’s ADRC in February 2024 following internal and external peer review (Awardee: JG, ADRC P30 grant, PI: Thomas J. Grabowski, MD). Phase One of Tool Development commenced February 21^st^, 2024. The results of the data analyses are expected to be available by August 2025.

### 3.1 Dissemination

The authors will disseminate the results of this multitiered work through academic and clinical conferences and peer-reviewed scientific journals. Results will also be disseminated via stakeholder forums, including the monthly online Memory and Brain Wellness newsletter and PPA Together! Support group. The authors will also develop opportunities to disseminate outcomes of the study to people living with PPA, SLPs, and researchers in collaboration with the National Aphasia Association PPA Task Force and supported by the International SLT/P PPA Network.

## 4 Discussion

To date, there are no standard training materials that enable SLPs to develop clinical expertise in PPA, nor frameworks to communicate assessment findings across health professions, particularly as they pertain to FC ability. In this project, we propose to develop a composite measure of FC that is structured as a simple rating scale and allows clinicians to use a common framework to synthesize speech, language, and communication function – regardless of the exact measurement tools used. Our aims are two-fold, to improve both clinical and research practices of PPA assessment. Clear and consistent agreement in behavioral ratings is paramount for appropriate clinical trial recruitment, the implementation of therapeutic intervention, and monitoring change over time. We will develop a series of tools that serve to train clinicians to assess PPA in speakers from an informative participant sample, and to create a validated assessment procedure to assess functional communication, which in turn provides the basis for researcher, practitioner, patient, and care partner education. The outcomes of this work will result in novel educational tools to cultivate comprehensive and resilience-oriented assessment processes, as well as stakeholder tools to provide direct feedback to PwPPA. Moreover, validation of the FC assessment will enhance collaboration and partnership amongst healthcare providers and across institutions that serve this population.

## Data Availability

No datasets were generated or analysed during the current study. All relevant data from this study will be made available upon study completion.

## 5 Acknowledgements

This work is supported by the generous University of Washington’s Alzheimer’s Disease Research Center Development Project Award (Awardee: JG).

## 6 Author Contributions

***Conceptualization:*** Jeanne Gallée, Paul K. Crane, Kimiko Domoto-Reilly, Rob J. Fredericksen

***Data curation:*** Jeanne Gallée, Paul K. Crane, Kimiko Domoto-Reilly, Rob J. Fredericksen

*Funding acquisition:* Jeanne Gallée

***Methodology:*** Jeanne Gallée, Paul K. Crane, Kimiko Domoto-Reilly, Rob J. Fredericksen, Aimee Mooney, Jade Cartwright, Maya L. Henry, Brielle C. Stark, Anna Volkmer

***Investigation:*** Jeanne Gallée, Paul K. Crane, Kimiko Domoto-Reilly, Rob J. Fredericksen, Aimee Mooney, Jade Cartwright, Maya L. Henry, Brielle C. Stark, Anna Volkmer

***Formal Analysis:*** Jeanne Gallée, Paul K. Crane, Kimiko Domoto-Reilly, Rob J. Fredericksen

***Supervision:*** Jeanne Gallée, Paul K. Crane, Kimiko Domoto-Reilly, Rob J. Fredericksen

*Writing – original draft:* Jeanne Gallée

***Writing – review & editing:*** Jeanne Gallée, Paul K. Crane, Kimiko Domoto-Reilly, Rob J. Fredericksen, Aimee Mooney, Jade Cartwright, Maya L. Henry, Brielle C. Stark, Anna Volkmer

## References

1. What Is Frontotemporal Dementia, the Disease Bruce Willis Is Diagnosed With? | AFTD [Internet]. 2023 [cited 2024 Feb 20]. Available from: https://www.theaftd.org/posts/all-us-states/what-is-frontotemporal-dementia-the-disease-bruce-willis-is-diagnosed-with/

2. Gorno-Tempini ML, Hillis AE, Weintraub S, Kertesz A, Mendez M, Cappa SF, et al. Classification of primary progressive aphasia and its variants. Neurology. 2011 Mar 15;76(11):1006–14.

3. Marshall CR, Hardy CJD, Volkmer A, Russell LL, Bond RL, Fletcher PD, et al. Primary progressive aphasia: a clinical approach. J Neurol. 2018 Jun;265(6):1474–90.

4. Ruksenaite J, Volkmer A, Jiang J, Johnson JC, Marshall CR, Warren JD, et al. Primary Progressive Aphasia: Toward a Pathophysiological Synthesis. Curr Neurol Neurosci Rep. 2021 Feb 4;21(3):7.

5. Belder CRS, Marshall CR, Jiang J, Mazzeo S, Chokesuwattanaskul A, Rohrer JD, et al. Primary progressive aphasia: six questions in search of an answer. J Neurol. 2024 Feb;271(2):1028–46.

6. Ramsberger G, Rende B. Measuring transactional success in the conversation of people with aphasia. Aphasiology. 2002 Mar 1;16(3):337–53.

7. Doedens WJ, Meteyard L. What is Functional Communication? A Theoretical Framework for Real-World Communication Applied to Aphasia Rehabilitation. Neuropsychol Rev. 2022 Dec;32(4):937–73.

8. Volkmer A, Cartwright J, Ruggero L, Beales A, Gallée J, Grasso S, Henry M, Jokel R, Kindell J, Khayum R, Pozzebon M. Principles and philosophies for speech and language therapists working with people with primary progressive aphasia: An international expert consensus. Disability and Rehabilitation. 2023 Mar 13;45(6):1063–78.

9. Kim SK, Park M. Effectiveness of person-centered care on people with dementia: a systematic review and meta-analysis. Clinical Interventions in Aging. 2017 Feb 17;12:381–97.

10. Hardy CJD, Taylor-Rubin C, Taylor B, Harding E, Gonzalez AS, Jiang J, et al. Symptom-led staging for semantic and non-fluent/agrammatic variants of primary progressive aphasia. Alzheimer’s & Dementia. 2024;20(1):195–210.

11. Gallée J, Cartwright J, Volkmer A, Whitworth A, Hersh D. “Please Don’t Assess Him to Destruction”: The R.A.I.S.E. Assessment Framework for Primary Progressive Aphasia. Am J Speech Lang Pathol. 2023 Mar 9;32(2):391–410.

12. Fazio S, Pace D, Flinner J, Kallmyer B. The Fundamentals of Person-Centered Care for Individuals With Dementia. The Gerontologist. 2018 Jan 18;58(suppl_1):S10–9.

13. Krein L, Jeon YH, Amberber AM, Fethney J. The Assessment of Language and Communication in Dementia: A Synthesis of Evidence. Am J Geriatr Psychiatry. 2019 Apr;27(4):363–77.

14. Ho T, Whitworth A, Hersh D, Cartwright J. “They are dealing with people’s lives…”: Diagnostic and post-diagnostic healthcare experiences in primary progressive aphasia. International Journal of Speech-Language Pathology. 2023 May 4;25(3):449–61.

15. Suárez-González A, Cassani A, Gopalan R, Stott J, Savage S. When it is not primary progressive aphasia: A scoping review of spoken language impairment in other neurodegenerative dementias. Alzheimer’s & Dementia: Translational Research & Clinical Interventions. 2021;7(1):e12205.

16. COMET Initiative | Primary Progressive Aphasia: a Core Outcome Set for improving intervention research [Internet]. [cited 2024 Mar 11]. Available from: https://www.comet-initiative.org/Studies/Details/1871

17. Gallée J, Volkmer A. A Window Into Functional Communication: Leveraging Naturalistic Speech Samples in Primary Progressive Aphasia. Perspect ASHA SIGs. 2021 Aug 20;6(4):704–13.

18. Kirshner B, Guyatt G. A methodological framework for assessing health indices. J Chronic Dis. 1985;38(1):27–36.

19. Hersh D, Boud D. Reassessing assessment: what can post stroke aphasia assessment learn from research on assessment in education? Aphasiology. 2024 Jan 2;38(1):123–43.

20. Buntinx WH. Understanding disability: A strengths-based approach. The Oxford handbook of positive psychology and disability. 2013 Aug 22:7–18.

21. Patel N, Peterson KA, Ingram RU, Storey I, Cappa SF, Catricala E, et al. A ‘Mini Linguistic State Examination’ to classify primary progressive aphasia. Brain Communications. 2022 Apr 1;4(2):fcab299.

22. Matias-Guiu JA, Grasso SM. Primary progressive aphasia: in search of brief cognitive assessments. Brain Communications. 2022 Oct 1;4(5):fcac227.

23. Sapolsky D, Domoto-Reilly K, Dickerson BC. Use of the Progressive Aphasia Severity Scale (PASS) in monitoring speech and language status in PPA. Aphasiology. 2014 Sep 2;28(8–9):993–1003.

24. McGee JS, McElroy M, Meraz R, Myers DR. A qualitative analysis of virtues and strengths in persons living with early stage dementia informed by the values in action framework. Dementia. 2023 Jan 1;22(1):46–67.

25. Loizidou M, Brotherhood E, Harding E, Crutch S, Warren JD, Hardy CJD, et al. ‘Like going into a chocolate shop, blindfolded’: What do people with primary progressive aphasia want from speech and language therapy? International Journal of Language & Communication Disorders. 2023;58(3):737–55.

26. Jünger S, Payne SA, Brine J, Radbruch L, Brearley SG. Guidance on Conducting and REporting DElphi Studies (CREDES) in palliative care: Recommendations based on a methodological systematic review. Palliative medicine. 2017 Sep;31(8):684–706.

27. Nasa P, Jain R, Juneja D. Delphi methodology in healthcare research: How to decide its appropriateness. World J Methodol. 2021 Jul 20;11(4):116–29.

28. Beiderbeck D, Frevel N, von der Gracht HA, Schmidt SL, Schweitzer VM. Preparing, conducting, and analyzing Delphi surveys: Cross-disciplinary practices, new directions, and advancements. MethodsX. 2021 Jan 1;8:101401.

29. Qualtrics [Internet]. [cited 2024 Mar 4]. Qualtrics XM - Experience Management Software. Available from: https://www.qualtrics.com/

30. Mauksch S, von der Gracht HA, Gordon TJ. Who is an expert for foresight? A review of identification methods. Technological Forecasting and Social Change. 2020 May 1;154:119982.

31. Sociocultural Research Consultants LLC. (2023). Dedoose v.9.0.90. Retrieved from www.dedoose.com. www.dedoose.com

32. Corbin J, Strauss A. Grounded theory methodology. Handbook of qualitative research. 1994;17:273–85.

33. The International Speech and Language Therapy / Pathology Primary Progressive Aphasia Network [Internet]. [cited 2024 Feb 20]. Available from: https://speechtherapyppa.builtbyknights.com/

34. Morris JC. Clinical Dementia Rating: A Reliable and Valid Diagnostic and Staging Measure for Dementia of the Alzheimer Type. International Psychogeriatrics. 1997 Dec;9(S1):173–6.

35. Morris JC, Ernesto C, Schafer K, Coats M, Leon S, Sano M, Thal LJ, Woodbury P. The Alzheimer’s Disease Cooperative Study. Clinical Dementia Rating training and reliability in multi-center studies: The Alzheimer’s Disease Cooperative Study Experience. Neurology. 1997;48:1508–10.

36. Wilson SM, Eriksson DK, Schneck SM, Lucanie JM. A quick aphasia battery for efficient, reliable, and multidimensional assessment of language function. Jäncke L, editor. PLoS ONE. 2018 Feb 9;13(2):e0192773.

37. Swinburn K, Best W, Beeke S, Cruice M, Smith L, Pearce Willis E, et al. A concise patient reported outcome measure for people with aphasia: the aphasia impact questionnaire 21. Aphasiology. 2019 Sep 2;33(9):1035–60.

38. Ericcson KA, Simon HA. Protocol analysis: Verbal reports as data (Rev. ed.). Cambridge, MA: Bradford. 1993.

39. Wolcott MD, Lobczowski NG. Using cognitive interviews and think-aloud protocols to understand thought processes. Currents in Pharmacy Teaching and Learning. 2021 Feb 1;13(2):181–8.

40. ParticiPics – Aphasia Institute [Internet]. [cited 2024 Feb 20]. Available from: https://www.aphasia.ca/participics/

41. McHugh ML. Interrater reliability: the kappa statistic. Biochemia Medica. 2012 Oct 15;22(3):276–82.

